# Improved deep learning model for differentiating novel coronavirus pneumonia and influenza pneumonia

**DOI:** 10.1101/2020.03.24.20043117

**Authors:** Min Zhou, Yong Chen, Dexiang Yang, Yanping Xu, Weiwu Yao, Jingwen Huang, Xiaoyan Jin, Zilai Pan, Jingwen Tan, Lan Wang, Yihan Xia, Longkuan Zou, Xin Xu, Jingqi Wei, Mingxin Guan, Jianxing Feng, Huan Zhang, Jieming Qu

## Abstract

**Background:** Chest CT had high sensitivity in diagnosing novel coronavirus pneumonia (NCP) at early stage, giving it an advantage over nucleic acid detection in time of crisis. Deep learning was reported to discover intricate structures from clinical images and achieve expert-level performance in medical image analysis. To develop and validate an integrated deep learning framework on chest CT images for auto-detection of NCP, particularly focusing on differentiating NCP from influenza pneumonia (IP).

**Methods:** 35 confirmed NCP cases were consecutively enrolled as training set from 1138 suspected patients in three NCP designated hospitals together with 361 confirmed viral pneumonia patients from center one including 156 IP patients, from May, 2015 to February, 2020. The external validation set enrolled 57 NCP patients and 50 IP patients from eight centers.

**Results:** 96.6% of NCP lesions were larger than 1 cm and 76.8% were with intensity below –500 Hu, indicating less consolidation than IP lesions which had nodules ranging 5-10 mm. The classification schemes accurately distinguished NCP and IP lesions with area under the receiver operating characteristic curve (AUC) above 0.93. The Trinary scheme was more device-independent and consistent with specialists than the Plain scheme, which achieved a F1 score of 0.847, higher than the Plain scheme (0.774), specialists (0.785) and residents (0.644).

**Conclusions:** Our study potentially provides an accurate early diagnosis tool on chest CT for NCP with high transferability, and shows high efficiency in differentiating NCP and IP, helping to reduce misdiagnosis and contain the pandemic transmission.

## Introduction

The spread of novel coronavirus pneumonia (NCP) induced by severe acute respiratory syndrome coronavirus 2 (SARS-CoV-2) has now entered a new phase in which new confirmed cases continue to decline in China while the novel virus rapidly spreading across the world with infected patients surging in hot spots of countries, such as European and Eastern Mediterranean region.

Early diagnosis is critical for both epidemic control and prompt medical intervention. Notably, pneumonia identified outside high-incidence areas are likely to be induced by a broad spectrum of pathogens, especially influenza which has high incidence in winter and spring. Influenza pneumonia (IP) brings huge burden to healthcare system due to its high morbidity and mortality rate. In United States, influenza accounted for more than 29 million infections and 16,000 deaths in 2019 (1). It was reported that oral oseltamivir accelerates symptom alleviation and reduces risks of lower respiratory tract complications in influenza (2). Therefore, early diagnosis and separation of IP patients from NCP patients will improve prognosis and optimize the allocation of medical resources. However, apart from overlapping symptoms and laboratory abnormalities, IP and NCP manifest similar chest CT findings (3), making it difficult to differentiate these two kinds of viral pneumonia.

The diagnostic efficiency of nucleic acid detection (4) is constrained by following limitations: 1) high false negative rate owing to low virus load at early infection stage (5) or possible genetic mutations (6); 2) shortage of detection reagents; and 3) long waiting time. It is found that some early-onset NCP patients who had already presented abnormal chest CT findings still got negative results on the initial nucleic acid test. As a result, the category of clinically diagnosed NCP was added in the fifth version of diagnosis and treatment scheme released by Chinese National Health

Commission (7), referring to suspected cases showing characteristics of viral pneumonia on chest CT, with the intention to reduce mortality rate and occurrence of cross infection while patients wait for laboratory confirmation. Additionally, CT has the advantage of evaluating the severity and surveilling the dynamic progress of pneumonia (8). The key issues in improving the capability to distinguish NCP from IP on chest CT scan are how to find the lesions quickly and make accurate differential diagnosis.

The problem could be alleviated by deep learning, a technique that has witnessed striking advances in healthcare applications (9, 10). It could achieve expert-level performance in medical image analysis with minimal time and labor cost, like detection of diabetic retinopathy and classification of skin cancer (11, 12). Deep learning is also widely used to automatically detect pneumonia based on chest X-ray images (13, 14), and discriminate usual interstitial pneumonia from nonspecific interstitial pneumonia based on chest CT images (15).

In this study, we developed and validated an integrated deep learning framework on chest CT images for auto-detection of NCP, particularly focusing on differentiating NCP from IP, ensuring prompt implementation of isolation. To alleviate transferability problem that a well-trained deep learning model performs poorly on data from unseen sources (16), we proposed a novel training scheme (Trinary scheme) to encourage the model to learn device independent features.

## Materials and methods

### 1. Patients

This retrospective study was conducted in eight tertiary referral centers (Center 1~8). In three designated hospitals for NCP screening, 35 confirmed NCP cases were enrolled as training set from a consecutive case series of 1138 suspected patients, together with 361 patients with confirmed viral pneumonia from Center one including 156 IP patients. Patients were diagnosed as viral pneumonia according to the 2016 Clinical Practice guidelines by the Chinese Thoracic Society and 2007 Infectious Diseases Society of America/American Thoracic Society guidelines (17, 18). The external validation set enrolled 57 NCP patients and 50 IP patients from eight hospitals. Inclusion and exclusion criteria, distribution of patients and flow chart of this study was shown in Figure 1. More details were in the Method E1.

**Figure 1.**
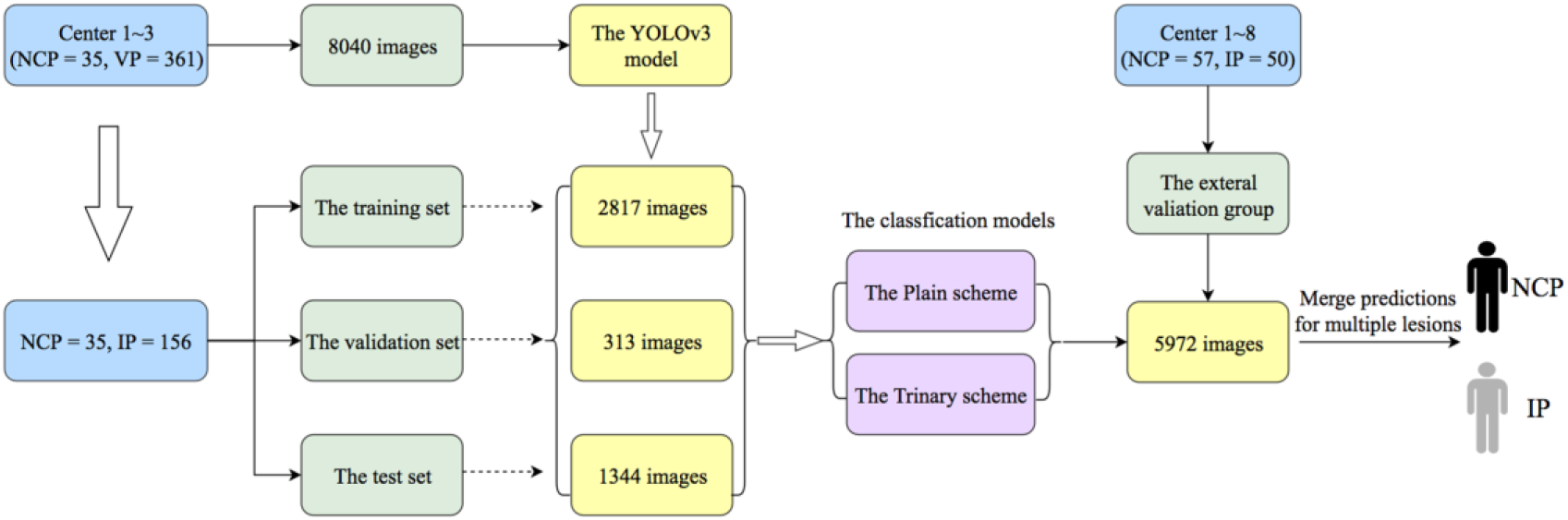
Flowchart Illustrating Deep Learning Process for Differentiating Diagnosis of NCP and IP from Multicenter. VP=viral pneumonia. NCP=novel coronavirus pneumonia. IP=influenza pneumonia.

### 2. Lesion detection

In our study, the lesion regions of each CT image were annotated by two radiologists, who have more than 10 years of experience in pulmonary-thoracic disease and were aware of the clinical history of infection. We used YOLOv3 to perform lesion detection on the selected images (19). The structure of YOLOv3 was presented on Figure E1 and detail information and CT slice thickness were presented on Table E1 and Method E2.

### 3. Lesion level classification

Because of the limited number of annotations, we chose VGGNet as the classification model (20). It is improved on the basis of AlexNet. To better fit our problem, we made some modifications on the original VGGNet and used transfer learning (16, 21). based on previous reports. We denoted the normal training process as the Plain scheme. To better solve the transferability problem of deep learning, we proposed a possible device-specific solution, named as the Trinary training scheme. The process for the Trinary scheme was described in the Method E3.

### 4. Patient level classification

Patient level classification was based on lesion level classification results. By taking sum of the predicted probabilities for all the lesions of a patient and then normalized between NCP and IP, we got the patient level classification. This simple averaging step could be considered as a model ensemble (22) for patient level classification.

### 5. Comparison with expert performance on the external validation set

To compare the performance of the deep learning framework and radiologists in the external validation group, a panel of ten radiologists were recruited. They were instructed to independently provide a classification decision on NCP or IP each time. We also classified the lesion by radiologists to determine which scheme was closer to the judgement of human experts. Details on evaluation were shown in the Method E4.

To better understand the performance difference between the Plain scheme and the Trinary scheme in different CT devices, we divided the NCP data on the external validation set into two categories. The first category contained 20 cases from centers (Center 1~3) that also appeared in the training set. The second category contained the remaining 37 cases from Center 4~8. We compared the performance between two schemes on the two categories.

### 6. Statistics

The classification metrics used included area under the receiver operating characteristic curve (AUC), sensitivity, specificity, accuracy, precision and F1 score. Details of statistics were in the Method E5.

## Results

### 1. Patient information

Details of clinical information for patients in the training, validation, test and external validation set were shown in Table E2 and the Result E1.

### 2. Comparison of imaging features between NCP and IP on the training and test set

We further performed a joint analysis of imaging features for the 35 NCP patients and 156 IP patients with 499 (1178) NCP (IP) lesions. 96.6% of NCP lesions were larger than 10 mm and 35.3% of the lesions were inhomogeneous, which was significantly different from that of IP (p=0.0094). Lesions with intensity less than - 500Hu accounted for 76.8% of lesions in NCP indicating less consolidation than IP. 5.4% lesions in IP were nodules (Hu>0) and 21 (5.6%) nodules of IP were 5-10 mm. Detailed information was presented on the Result E2, Table E3 and Figure E2-E3. Examples for CT image features of NCP and IP patients were shown on Figure 2 and Figure E4.

**Figure 2.**
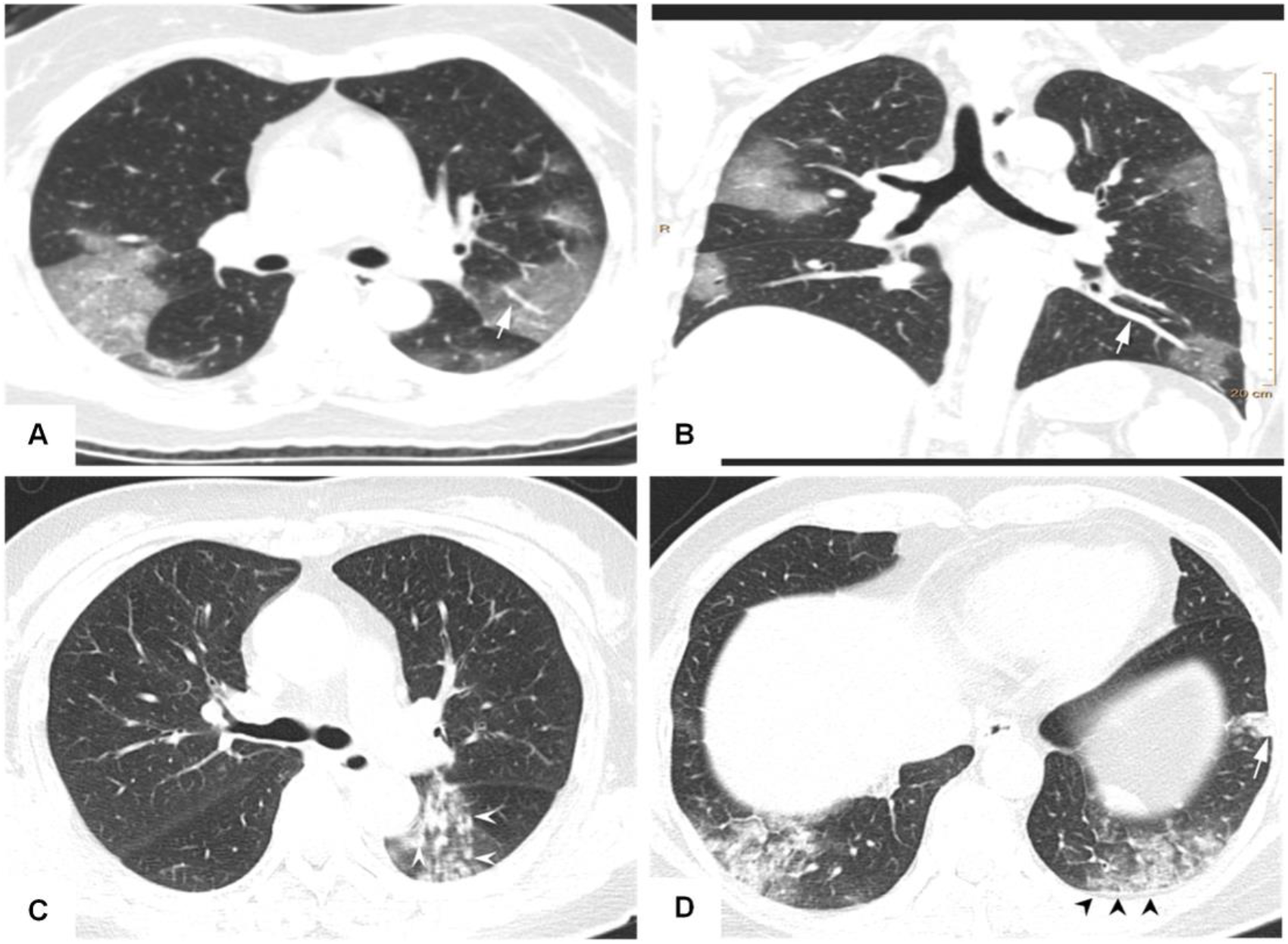
CT Image Features of an NCP Patient and an IP Patient. (A-B) Chest CT of an NCP patient: 45-year-old female with history of stay in Wuhan for two days. Presenting with fever and cough for 4 days, she was proved to have the NCP. CT scan (A) shows bilateral GGO scatted in four lobes with an obvious peripheral distribution and bilateral lobular or subsegmental GGO involving mainly the subpleural lung regions. Vascular dilation with GGO surrounding was more evident and pulmonary venous branch passed through the lesion with luminal dilation (white arrow) and it is more obvious in maximum-intensity projection (MIP) imaging (B). (C-D) CT images of an IP patient: CT scan shows multiple, bilateral, and randomly distributed small ill-defined nodules (white arrow head) with small branch opacities indicating the bronchiolitis. Peripheral subpleural consolidation (white arrow) in the left lower lobe with the interlobular septum and pleura thickening (black head).

### 3. Lesions could be effectively detected and classified by deep learning models

We applied the lesion detection model on the test set. The detected lesion examples were presented on Figure E5. The results showed that the detection performance was not sensitive to the confidence score as long as the cutoff for confidence score was in a reasonable range (Table E4). The detection model achieved F1 score 0.742 under confidence cutoff 0.1. We further used the annotated lesions to train and evaluate the model. Trinary scheme (with AUC 0.95) performed better than the Plain scheme (with AUC 0.93) (Figure E6). More performance measures can be found in Table E5. Two experienced specialists classified the lesions on which two schemes made very different predictions (with probability difference no less than 0.5) (Figure E7). 366 (or 174) out of 540 NCP lesions were identified by Trinary (or Plain) scheme correctly. Detailed analysis showed that the Plain scheme tend to yield unreasonably high or low probability of lesion predictions depending on the lesions from centers in the training set or not. The results indicated the Trinary scheme was more consistent with specialists than Plain scheme on the lesion level classification. Detailed information is presented in the Results E3-E5.

### 4. Trinary scheme outperformed the specialist group on patient level classification

The performance of human experts for patient classification was shown in Table E6 and the Result E6. Both of the specialist group and the resident group reached good consistency, with intraclass correlation coefficient (ICC) of 0.899 and 0.798, respectively. Correlation for 10 radiologists was presented on Table E7.

For the Plain and Trinary scheme, it took 10 seconds to detect and classify all detected lesions for a single patient on average. Figure 3 showed the ROC curves for the test and the external validation set of both training schemes. On the test set, the Plain and Trinary scheme performed similarly good with AUC of 0.99 (Figure 3A). The AUCs were much higher than the AUCs on the lesion level owing to the ensemble effect. For both schemes, the sensitivity is 100%. The specificities were 92.5% and 95% for the Plain and Trinary scheme, respectively (Table E8). On the external validation set, 13 (22.8%) patients with NCP were correctly classified by our Trinary scheme, but misdiagnosed by at least three specialists. Among them, 4 patients had the CT findings less frequently reported in other NCP cases, such as a small mixed ground glass opacity in the central part or solitary consolidation. They were misdiagnosed by five specialists (Figure 4). The results indicated both of the schemes surpassed the discrimination capability for residents for NCP and IP and achieved specialist level. More details were shown in the Result E7.

**Figure 3.**
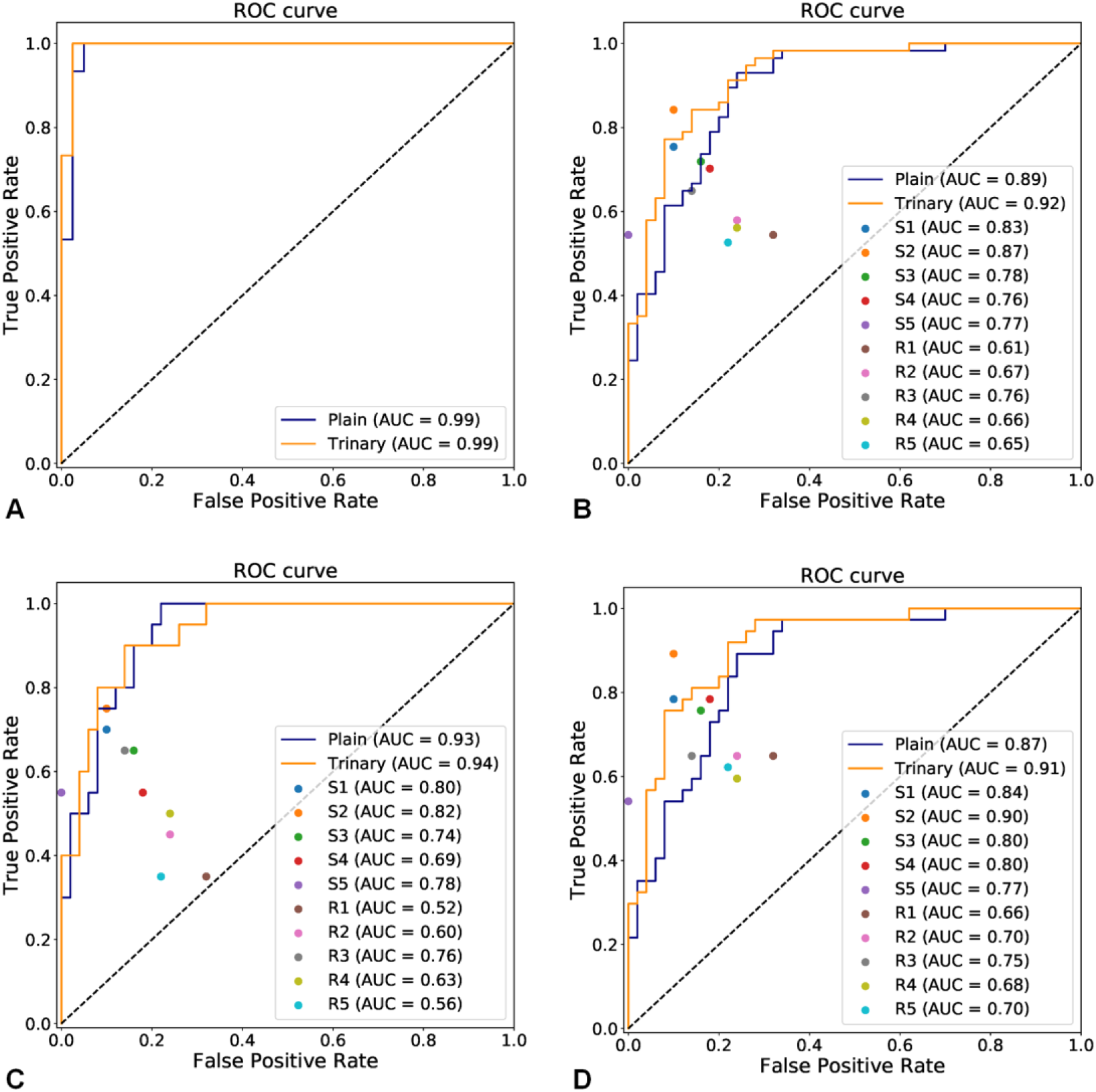
Receiver Operating Characteristic Curves of Patient Level Classification. (A) both of the schemes achieved an AUC of 0.99 for differentiating diagnosis of NCP and IP patients on the test set. (B) AUCs of deep learning schemes and human experts for differentiating diagnosis on the external validation set. Both of the schemes performed better than most of the human experts and the Trinary scheme (AUC 0.92) performed better than the Plain scheme (AUC 0.89). Specialist group (S1~S5) performed better than the resident group (R1~R5) did. (C) AUCs on patients in the first category (all 50 IP cases and 20 NCP patients from Center 1~3). The performance of both schemes is very similar (AUC 0.93 and 0.94). (D) AUCs on patients in the second category (all 50 IP cases and 37 NCP cases from Center 4~8). The performance of Trinary schemes is better than plain scheme (AUC 0.91 and 0.87).

**Figure 4.**
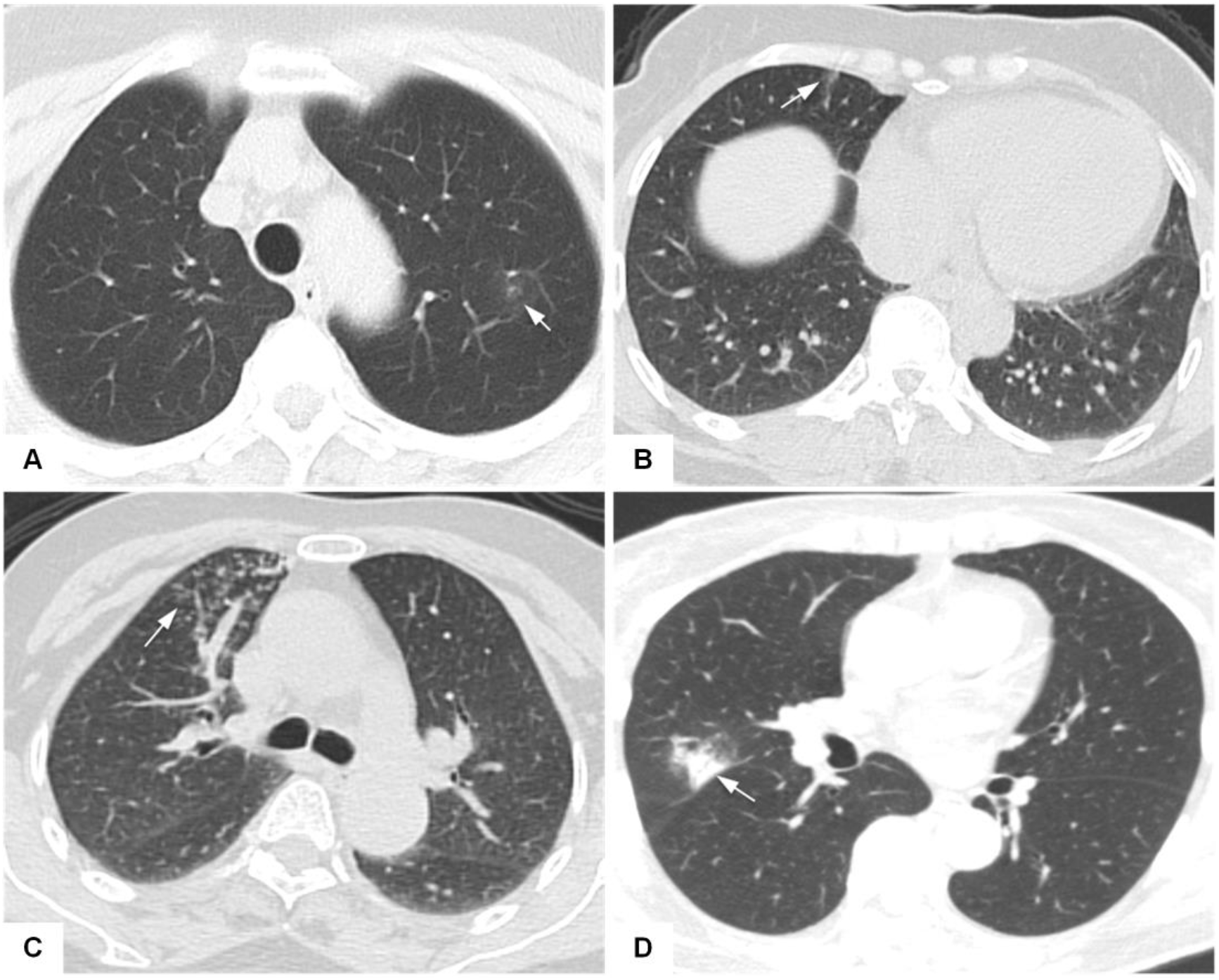
CT Images of Four NCP Patients Misdiagnosed by Specialists but Correctly Classified by Trinary Schemes. Axial CT plain scan in patients suffering from NCP which were misdiagnosed as IP by the specialist panel. (A). 50-year-old female with small piece of pure GGO in left upper lobe (arrow); she was negative in nucleic acid tests twice until positive in the third time (predicted probability 0.573); (B). 49-year-old female also with a small GGO (arrow) in right middle lobe (predicted probability 0.56); (C). 65-year-old female with patchy and punctate GGO and small nodules (arrow) in right middle lobe (predicted probability 0.515); (D). 65-year-old female with patchy consolidation (arrow) (predicted probability 0.831). Their common characteristics were either small lesions located in one segment or lack of specificity of typical findings in NCP. Patients above were correctly classified as NCP by Trinary scheme. (Predicted probability represented the probabilities of being NCP by the Trinary scheme)

As expected, both the Plain and the Trinary scheme performed better on cases from centers if the training data contained cases from same centers. Both schemes performed similarly (AUC 0.93 and 0.94, Figure 3C). Importantly, Trinary scheme performed better (AUC 0.91) than Plain scheme (AUC 0.87) on the second category (data was not from centers included in the training set) (Figure 3D). In terms of F1 measure, the Trinary scheme achieves score 0.847, which is higher than the Plain scheme (0.774) and also much higher than the specialist group (average 0.785) and the resident group (average 0.644). Trinary scheme was better correlated with specialists in both categories (Table E9). More details were in the Result E7.

### 5. Trinary scheme performed better on new CT devices

Table 1 summarized the CT devices on which both schemes and ten radiologists made wrong classification on cases from the external validation set. We first observed that the IP cases are from 10 CT devices, despite the fact that they were from the same center. The majority of the tested IP cases have been correctly classified by both schemes. The only exception was uCT 528, a new CT device. On uCT 528, eight patients were examined, seven of them were IP from Center 1, from which six and five cases were misclassified by Plain and Trinary scheme respectively. Yet more than three patients were also misdiagnosed by the specialists. The main manifestations were peripheral single or multiple ground grass opacities with or without patchy consolidation in the lower lobe or bilateral distribution, which mimic the findings of NCP, leading to the misclassification (Figure E8). Another one was from Center 8 which was an NCP but misdiagnosed by all specialists (Figure 4A). The Trinary scheme performs better than the Plain scheme in this situation.

**Table 1.**
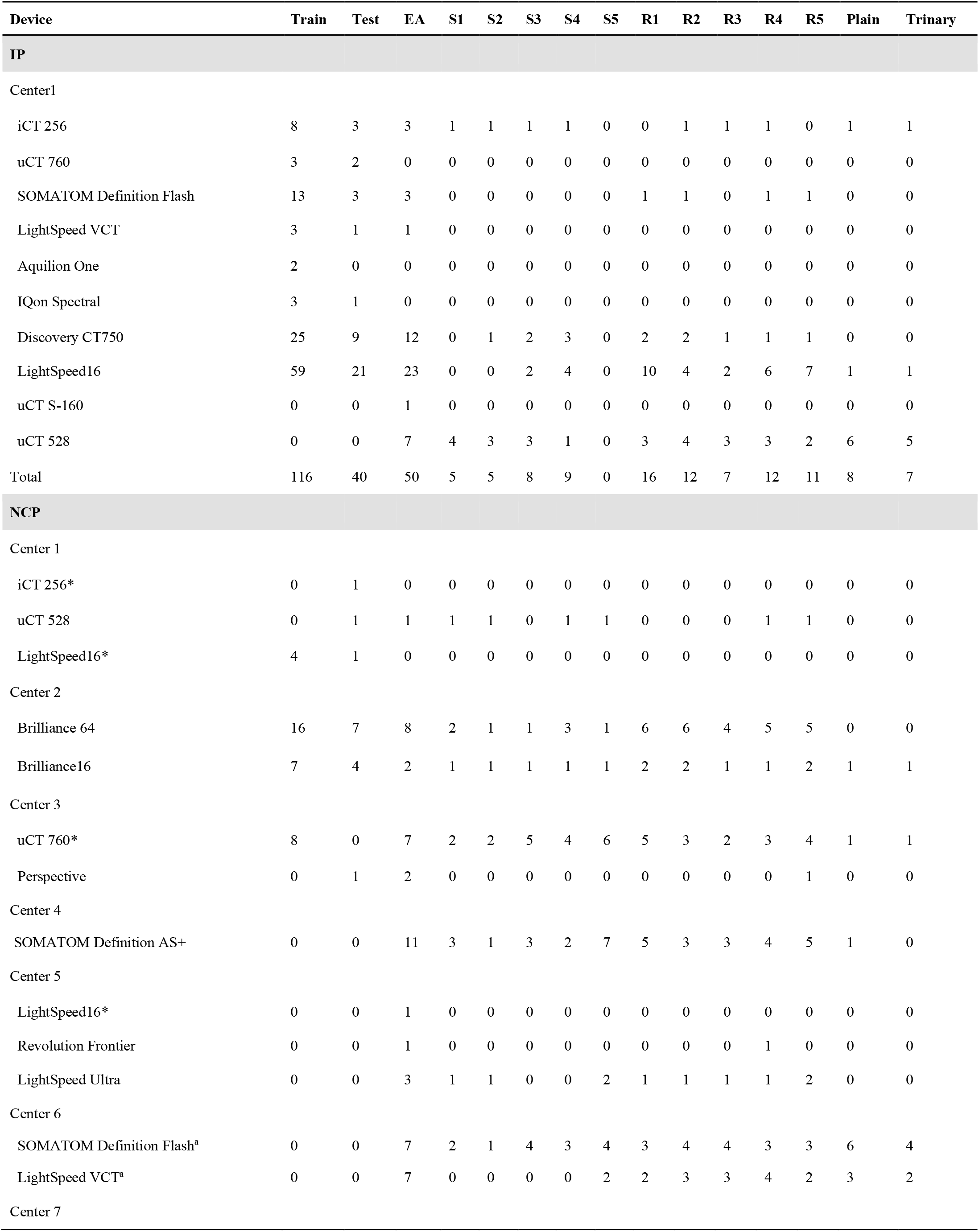
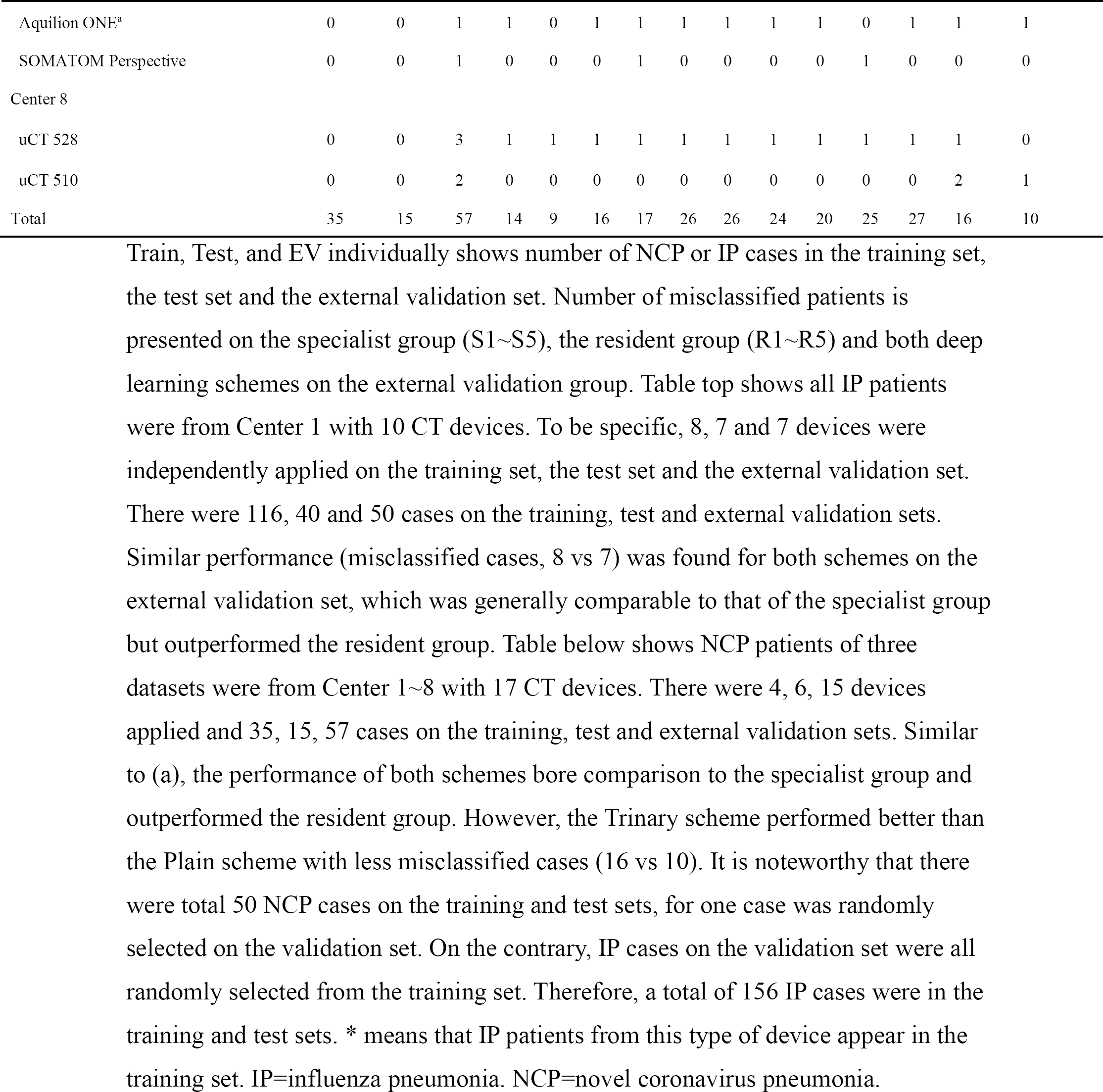
Statistics Specific to Each Center and Device on the External Validation Set.

As shown in Table 1, the Plain scheme misclassified 16 cases and the Trinary scheme reduced it to 10. The error rates of both schemes for the two CT devices (SOMATOM Definition Flash and LightSpeed VCT) from Center 6 were exceptionally high compared to all other CT devices. As both of them only contributed IP training cases for the model, the classification model may learn the device specific features, and wrongly treated these features as specific to IP during training. During testing, the schemes would therefore tend to wrongly classify NCP.

Similar problems have been observed in previous study (23). The Trinary scheme performed better than the Plain scheme on these devices, implying that the Trinary scheme is less influenced by the device specific features. Detailed information was in the Results E8.

## Discussion

The escalating crisis caused by SARS-CoV-2 with high infectivity and multiple routes of transmission is complicated by its co-occurrence with seasonal influenza, exactly as the things happening in the United States that some COVID-19 deaths have been misdiagnosed as influenza. The similarities in clinical symptoms between NCP and IP, along with shortage and high false negative rate of nucleic acid detection kits, make the differential diagnosis difficult (24–26), prompting clinicians to investigate new diagnostic methods. Chest CT had high sensitivity in diagnosing NCP at early stage, giving it an advantage over nucleic acid detection in time of crisis. This is the reason why Hubei Provincial Government adopted characteristic chest CT finding as an important criterion for diagnosis of NCP at the peak of outbreak. However, the similar chest CT manifestations of NCP and IP will inevitably lead to inaccurate diagnosis even for experienced physicians, and increase the risk of over-diagnosis and cross infection (3, 8). The main challenge to employ CT as a predominant diagnostic tool is to improve the accuracy and speed in identifying specific lesions on chest CT images.

We first annotated NCP and IP lesions and analyzed the difference of their chest CT features. We found that 76.8% of lesions in NCP are less than -500 HU, 96.1% of NCP patients had bilateral lung damage and 33.3% had all five lung lobes affected, consistent with the pathophysiology of NCP. SARS-CoV-2 is presumed to bind to angiotensin converting enzyme 2 (ACE2) receptor (27) concentrated on alveolar type-2 epithelial cells, which will undergo apoptosis after infection, leading to diffuse alveolar damage and interstitial fluid absorption disorder (28). Pathological findings of NCP showed pulmonary edema and hyaline membrane formation (29). While influenza viruses primarily cause damage to the trachea epithelial cells, leading to necrotizing bronchitis and diffuse alveolar damage to the upper respiratory tract (3). It was reported that the size of the nodules helps to differentiate different types of infections, for instance, the nodules of viral infection are ordinarily less than 10 mm (30). Consistent with previous reports, we found that nodules are present more often in IP with their sizes ranging 5-10 mm.

Based on above observations, we constructed an integrated artificial intelligence (AI) framework consisting of two deep learning models. The YOLOv3 model is applied to identify lesions, followed by lesion classification by the modified VGGNet. During developing the deep learning model, the first problem we met lies in transferability (5, 6). The model performs better on cases from CT device appearing in the training set than cases from CT devices not included. To address this problem, especially when classifying image data from multiple CT devices, we proposed a Trinary classification scheme to penalize the network from extracting device specific features during learning. By doing so, it would lead to high cost on the random region inputs, forcing the model to extract more lesion specific features. Although it is impossible to exclude all device specific features, we observed a visible improvement in performance (AUC from 0.85 to 0.89) on patient level classification. Such a performance is comparable with the judgement of experienced specialists. 13 (22.8%) NCP patients presenting uncommon CT findings, such as a small ground-glass opacity (GGO) in the central part were correctly classified by our Trinary scheme, instead, were misdiagnosed by three specialists. We have verified the clinical applicability of our developed AI model by including data from multiple machines and centers. We first demonstrated that the AI model performed well using training and test data from four machines of three centers with an AUC of 0.99. Similar performance of the model with specialists on independent verification data from fifteen machines of eight centers further suggests good clinical applicability.

Although our AI system achieved good performance, it misclassified a small number of NCP and IP patients, which may be caused by poor spatial resolution of some of images. In this study, we used 5 mm instead of 1 mm layer thickness in CT reconstruction, which would limit our capability to detect small lesions. Nevertheless, 5 mm layer thickness is a standard parameter in most hospitals and is sufficient to identify major imaging differences between NCP and IP as demonstrated by our study. Therefore, it is worthy to sacrifice certain accuracy to provide wider applicability of the deep learning model.

Currently, SARS-CoV-2 is wildly spreading around the world, efficient and accurate diagnosis of NCP is crucial for prevention and control. Our deep learning model potentially provides an accurate early diagnostic tool for NCP, especially when nucleic acid test kits are short of supply, which is a common problem during outbreaks. This could help reduce the missed diagnosis rate and diagnosis time, ensure prompt patient isolation and early treatment, improve prognosis and largely prevent transmission. The high efficiency of our model to differentiate NCP and IP could be very beneficial to reduce misdiagnosis rate and optimize the allocation of medical resources, particularly in areas with high prevalence of both NCP and IP. Trinary scheme not only improves the performance of the model in discriminating NCP from IP, but also behaves more similar to specialists than the Plain scheme. Because the proposed Trinary scheme is designed for general purpose, we believe that it can be applied to a wide range of medical image classification.

## Data Availability

Due to patient privacy concerns the datasets are currently not publicly available. However, the datasets analyzed during the current study are available from the corresponding author on reasonable request.

## Acknowledgements

We would like to thank all the radiologists who helped with the analysis and interpretation of the imaging data.

